# COVID-19 is associated with higher risk of venous thrombosis, but not arterial thrombosis, compared with influenza: Insights from a large US cohort

**DOI:** 10.1101/2021.10.15.21264137

**Authors:** Andrew Ward, Ashish Sarraju, Donghyun Lee, Kanchan Bhasin, Sanchit Gad, Rob Beetel, Stella Chang, Mac Bonafede, Fatima Rodriguez, Rajesh Dash

## Abstract

**Introduction:** Infection with SARS-CoV-2 is typically compared with influenza to contextualize its health risks. SARS-CoV-2 has been linked with coagulation disturbances including arterial thrombosis, leading to considerable interest in antithrombotic therapy for Coronavirus Disease 2019 (COVID-19). However, the independent thromboembolic risk of SARS-CoV-2 infection compared with influenza remains incompletely understood. We evaluated the adjusted risks of thromboembolic events after a diagnosis of COVID-19 compared with influenza in a large retrospective cohort.

**Methods:** We used a US-based electronic health record (EHR) dataset linked with insurance claims to identify adults diagnosed with COVID-19 between April 1, 2020 and October 31, 2020. We identified influenza patients diagnosed between October 1, 2018 and April 31, 2019. Primary outcomes [venous composite of pulmonary embolism (PE) and acute deep vein thrombosis (DVT); arterial composite of ischemic stroke and myocardial infarction (MI)] and secondary outcomes were assessed 90 days post-diagnosis. Propensity scores (PS) were calculated using demographic, clinical, and medication variables. PS-adjusted hazard ratios (HRs) were calculated using Cox proportional hazards regression.

**Results:** There were 417,975 COVID-19 patients (median age 57y, 61% women), and 345,934 influenza patients (median age 47y, 66% women). Compared with influenza, patients with COVID-19 had higher venous thromboembolic risk (HR 1.53, 95% CI 1.38–1.70), but not arterial thromboembolic risk (HR 1.02, 95% CI 0.95–1.10). Secondary analyses demonstrated similar risk for ischemic stroke (HR 1.11, 95% CI 0.98–1.25) and MI (HR 0.93, 95% CI 0.85–1.03) and higher risk for DVT (HR 1.36, 95% CI 1.19–1.56) and PE (HR 1.82, 95% CI 1.57–2.10) in patients with COVID-19.

**Conclusion:** In a large retrospective US cohort, COVID-19 was independently associated with higher 90-day risk for venous thrombosis, but not arterial thrombosis, as compared with influenza. These findings may inform crucial knowledge gaps regarding the specific thromboembolic risks of COVID-19.

## INTRODUCTION

Severe acute respiratory syndrome coronavirus 2 (SAR-CoV-2) is typically compared with the influenza virus in literature to help contextualize its natural history and risks [1,2]. While SARS-CoV-2 and influenza have certain overlapping characteristics (such as fever syndromes and respiratory droplet spread), there are several clinical features and outcomes that are potentially distinct to coronavirus disease 2019 (COVID-19). In particular, prior work has linked SARS-CoV-2 infection with coagulation disturbances and elevated thromboembolic risk including arterial events such as ischemic stroke [3]. This has led to substantial contemporary interest in the role of thromboprophylaxis in COVID-19 management [4,5]. Health systems implemented antithrombotic strategies for COVID-19 during the pandemic, including treatment-dose anticoagulation [6], and major clinical trials of prophylactic anticoagulation in COVID-19 were initiated [4,7].

Data linking SARS-CoV-2 and thromboembolic events were initially derived from real-world COVID-19 case series or single-arm cohorts suggesting high thromboembolic event rates in patients with COVID-19 [3,8–10]. A cohort study of patients in New York City that compared COVID-19 with influenza further suggested that patients with COVID-19 who were in the emergency department or were hospitalized had higher rates of ischemic stroke compared with patients who were in the emergency department or hospitalized with influenza [11]. This analysis was, however, limited by the incorporation of only a few selected covariates and a lack of broad adjustment for baseline cohort characteristics that may affect thromboembolic outcomes. Robust comparative data regarding the independent risk of arterial and venous thromboembolic events in COVID-19 versus influenza remain lacking. Accurately assessing the thromboembolic risks of SARS-CoV-2 infection is essential to inform the potential benefits and risks of thromboprophylaxis in COVID-19 and guide future studies.

Despite high interest, the independent risk of thromboembolic events with SARS-CoV-2 infection compared with influenza after accounting for baseline cohort characteristics remains incompletely understood. To address such crucial COVID-19 knowledge gaps, the FDA Sentinel System for COVID-19 activities aims to promote timely research efforts [12]. As a part of the COVID-19 Evidence Accelerator convened by the Reagan-Udall Foundation for the FDA, linked with the FDA Sentinel System, we evaluated the independent risk of arterial and venous thrombotic events in patients diagnosed with COVID-19 compared with patients diagnosed with influenza in a large retrospective cohort, after accounting for baseline cohort characteristics. We hypothesized that a diagnosis of COVID-19 would be associated with elevated arterial and venous thromboembolic risk compared with influenza.

## METHODS

### Study Design and Data Sources

We performed a retrospective cohort study on the Veradigm Health Insights Ambulatory EHR Research Database linked with insurance claims data from January 1, 2015 to October 31, 2020. The electronic health record dataset consists of de-identified patient records sourced from ambulatory/outpatient primary care and specialty settings. The insurance claims data contains a de-identified combination of closed and open claims from inpatient and outpatient locations.

This work was part of the COVID-19 Evidence Accelerator convened by the Reagan-Udall Foundation for the FDA, in collaboration with Friends of Cancer Research, which assembles experts from the health systems research, regulatory science, data science, and epidemiology to participate in parallel analyses focusing on coagulopathy in COVID-19. Analytic partners aligned on a common protocol (the current study’s methodology was adapted from the Sentinel Initiative for the FDA Natural History of Coagulopathy in COVID-19 Study Synopsis and Statistical Analysis Plan [12,13]) and conducted analyses independently; methods and results were shared side-by-side to evaluate differences and similarities. This methodology has been successfully applied to other COVID-19 research questions [14].

### Study Cohort

Two cohorts were defined: a COVID-19 cohort and a comparison influenza cohort. For the COVID-19 cohort, individuals over the age of 18 were included if, after April 1, 2020, they had a COVID-19 diagnosis based on International Classification of Diseases, Clinical Modification, 10th revision (ICD-10-CM) codes or molecular testing via Logical Observation Identifiers Names and Codes (LOINC) from an inpatient or outpatient setting (from either insurance claims or electronic health records, Supplemental Tables 1 & 2). Individuals over the age of 18 were included in the influenza cohort if, between October 1, 2018 and April 30, 2019, they had an influenza diagnosis based on ICD-10-CM codes or molecular testing from an inpatient or outpatient setting. Separate study years were chosen to minimize the possibility of influenza cohort patients having undiagnosed COVID-19. ICD-10-CM and LOINC lists were defined by the Sentinel Initiative [12].

The date of the first COVID-19/influenza diagnosis, or date of specimen collection of the first positive molecular test, served as the index date for each individual. Individuals were excluded if they 1) had evidence of COVID-19 in the 90 days prior to their index date; 2) did not have either A) at least two insurance claims in the 365 days on or before the index date or 365 days of continuous coverage as indicated by closed claims enrollment, or B) at least 2 encounters in the EHR in the 365 days on or before the index date; 3) were less than 18 years of age on their index date; 4) had evidence of coinfection with another respiratory virus (RSV, adenovirus, paraflu, enterovirus, rhinovirus, human metapneumovirus, and, for the COVID-19 cohort only, influenza) based on ICD-10-CM codes within 14 days of COVID-19/influenza diagnosis; 5) had no claims at any point after the index date.

### Data Collection

Comorbidities associated with cardiovascular disease and COVID-19 were collected during the baseline period, defined as the 365 days prior to the index date. The comorbidities included angina, atrial fibrillation, antiphospholipid antibody syndrome, alcohol abuse, cancer (excluding non-melanoma skin cancers), current tobacco use, current pregnancy, chronic kidney disease, chronic obstructive pulmonary disease, diabetes mellitus (type 1 and type 2), heart failure, HIV infection, hyperlipidemia, hypertension, inherited (primary) thrombophilia, polycythemia, thrombocytosis, neurological disease, obesity, prior ischemic stroke, prior MI, cardiovascular disease, cerebrovascular disease, peripheral arterial disease, and venous thromboembolism (i.e., deep venous thrombosis, pulmonary embolism, and/or thrombosis due to cardiac/vascular prosthetic device, implant, or graft).

Medication prescription fill data was collected from insurance claims. Medication usage was ascertained via the presence of a prescription fill 183 days to 3 days before index date (we excluded the three days before COVID-19 diagnosis to minimize protopathic bias). Medications assessed included anticoagulants, antiplatelets, and statins.

Diagnosis information was captured using ICD-10-CM codes. ICD code lists used to define relevant clinical concepts, NDC codes to define medication categories, and LOINC codes to define laboratory test results were leveraged from existing code lists from the Sentinel Initiative led by the FDA and HealthPals.[15]

### Outcomes

The two co-primary endpoints for this study were 1) a composite arterial thrombosis outcome, consisting of myocardial infarction (MI) and ischemic stroke; and 2) a composite venous thromboembolism outcome, consisting of acute deep vein thrombosis (DVT) and pulmonary embolism (PE). Endpoints were evaluated in the 90 days after diagnosis of COVID-19 or influenza, inclusive of the index date.

Outcomes were identified using ICD-10-CM codes from previously validated ICD diagnosis lists [16–20]). While MI, ischemic stroke, and PE were ascertained from ICD codes restricted to the inpatient setting, acute DVT was identified via ICD code from either the inpatient or emergency department setting since uncomplicated DVTs may be treated in the emergency department setting without hospitalization (see Supplement for more details).

Secondary arterial outcomes included MI and ischemic stroke (evaluated separately), and a composite of a broader range of arterial thrombotic complications: angina, transient ischemic attack, coronary angioplasty, coronary artery bypass grafting, peripheral artery disease, and amputation, and were identified via ICD codes and/or CPT/HCPCS codes from the inpatient or emergency department setting.

Secondary venous outcomes included acute DVT and PE (evaluated separately), and a composite outcome consisting of acute DVT, PE, and venous thrombosis of devices, implants, or grafts (identified via ICD code from the inpatient or emergency department setting).

### Study Endpoint

The study endpoint was the first of either 1) outcome, 2) 90 days after index date, 3) end of an individual’s data record (defined as either the last day on which an insurance claim was recorded or the last day on which an individual was enrolled in an insurance plan, whichever occurred later). Competing outcomes were not treated as censoring events; in other words, each outcome was evaluated separately. For example, while analyzing the primary arterial endpoint, a patient who experienced a DVT event in the follow-up period would continue to be monitored for an arterial event after their DVT.

### Statistical Analysis

Differences between the COVID-19 cohort and the influenza cohort were assessed using standardized differences. A standardized difference of ≥0.10 indicated meaningful imbalance [21]. The absolute risk and unadjusted incidence rates of the primary outcomes, secondary outcomes, and composite outcomes were calculated over the follow-up period of 90 days.

Propensity scores were used to control for confounding. Propensity scores were estimated using a logistic regression model with the group status (COVID-19/influenza) as the dependent variable. The following covariates were included in the regression model: age category, sex category, severity of infection, care setting of diagnosis, prior institutional stay, history of cardiovascular disease, venous thromboembolism, ischemic stroke, myocardial infarction, atrial fibrillation, heart failure, hyperlipidemia, hypertension, peripheral arterial disease, diabetes, neurologic disease that promotes stasis/immobility, obesity, chronic kidney disease, cancer, COPD, rheumatic disease, antiphospholipid antibody syndrome, inherited thrombophilia, polycythemia, thrombocytosis, current pregnancy, alcohol abuse, current tobacco use, and use of anticoagulants, antiplatelets, statins, oral chemotherapeutics, oral contraceptives, estrogen replacement, or testosterone replacement.

Individuals whose propensity score exceeded the maximum or minimum scores present in the other cohort were excluded (a method known as “trimming the tails”). Propensity score stratification, with sample average treatment effect weights, was used to assign a weight to each individual [22]. A total of 50 bins were used in the propensity score stratification process, spaced evenly from the highest to the lowest propensity score (after trimming the tails). More details are available in the supplement.

Weighted Cox proportional hazards regression, accounting for the propensity score, was used to calculate hazard ratios with robust 95% confidence intervals for the primary outcome and expanded set of secondary outcomes. Analysis was performed in Python 3.8.

### Sensitivity Analysis

Individuals were further stratified by prior cardiovascular disease (with/without), prior venous thromboembolism (with/without), or care setting of diagnosis (inpatient hospital or outpatient). Propensity scores and stratified weights were recalculated for each sensitivity cohort. Arterial outcomes (primary and secondary composite arterial endpoints, ischemic stroke, and MI) were analyzed after stratification by prior cardiovascular disease; venous outcomes (primary and secondary composite venous endpoints, acute DVT, and PE) were assessed when stratifying by prior VTE; and all outcomes were analyzed when stratifying by care setting of diagnosis.

## RESULTS

### Cohorts

There were 417,975 COVID-19 patients and 345,934 influenza patients who met study eligibility criteria (Figure 1). The COVID-19 cohort had a median age of 57 (interquartile range [IQR] 40-72) years, with 61% women (Table 1). The influenza cohort had a median age of 47 (IQR 32-61) years, with 66% women. A total of 194,346 (46%) COVID-19 patients and 252,001 (73%) influenza patients were diagnosed in an outpatient setting. A total of 225,988 (54%) COVID-19 patients and 142,789 (41%) influenza patients had prior cardiovascular disease, and 14,062 (3.4%) COVID-19 patients and 6,614 (1.9%) influenza patients had prior VTE.

**Figure 1:**
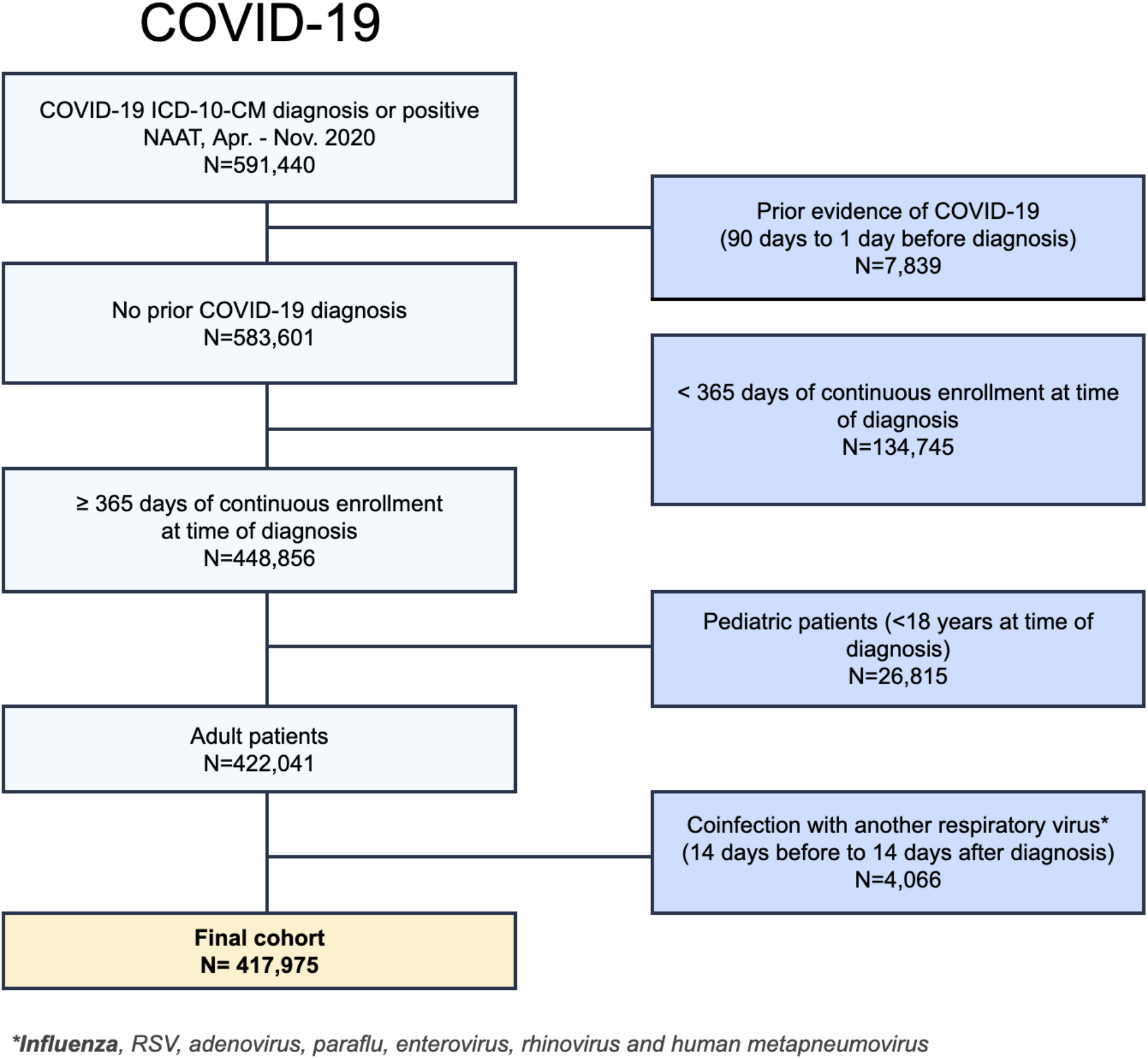
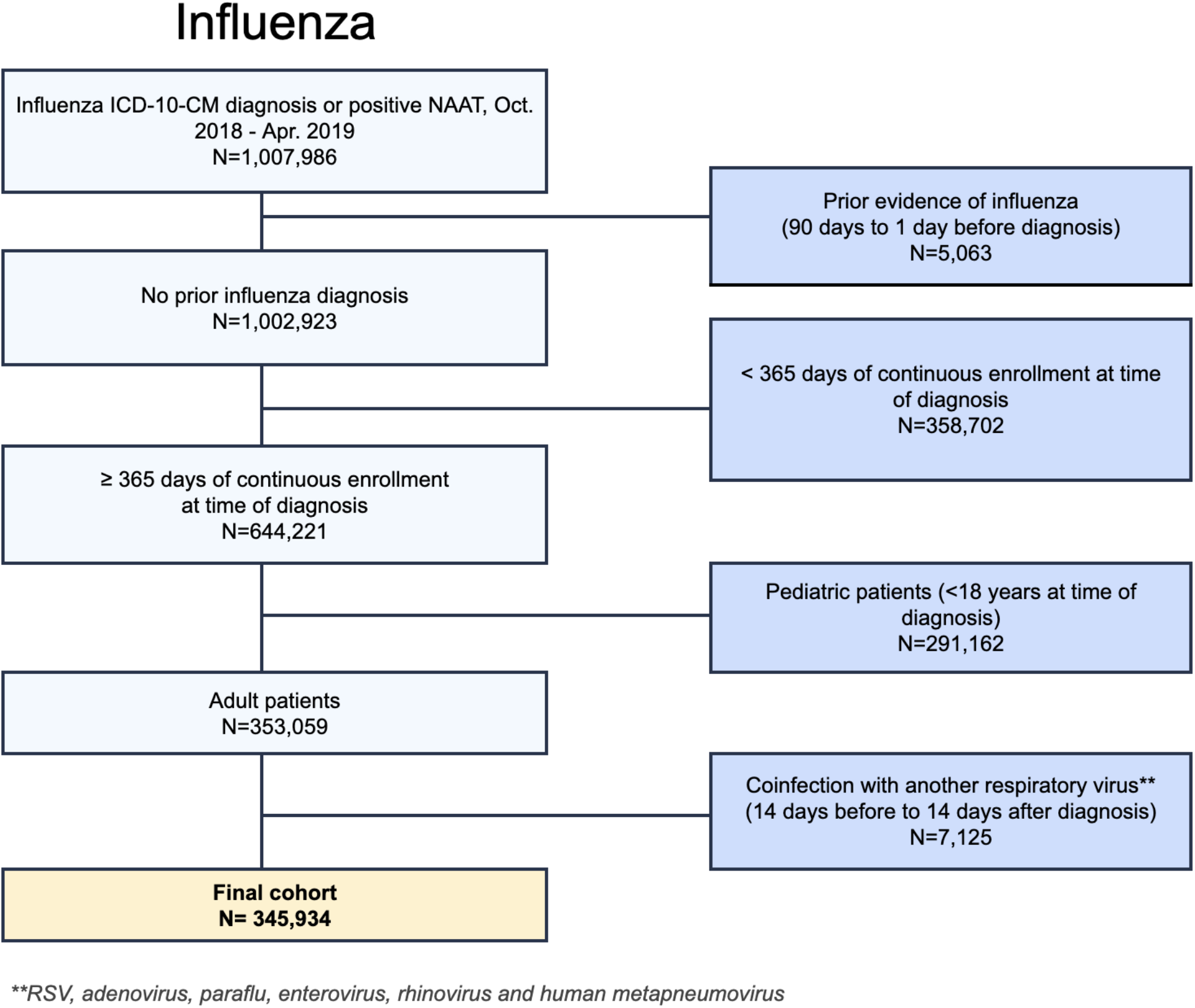
Cohort selection diagram for the COVID-19 cohort (a) and for the influenza cohort (b) Abbreviations: ICD-10-CM - international classification of diseases, 10th revision; NAAT - nucleic acid amplification test

**Table 1:**
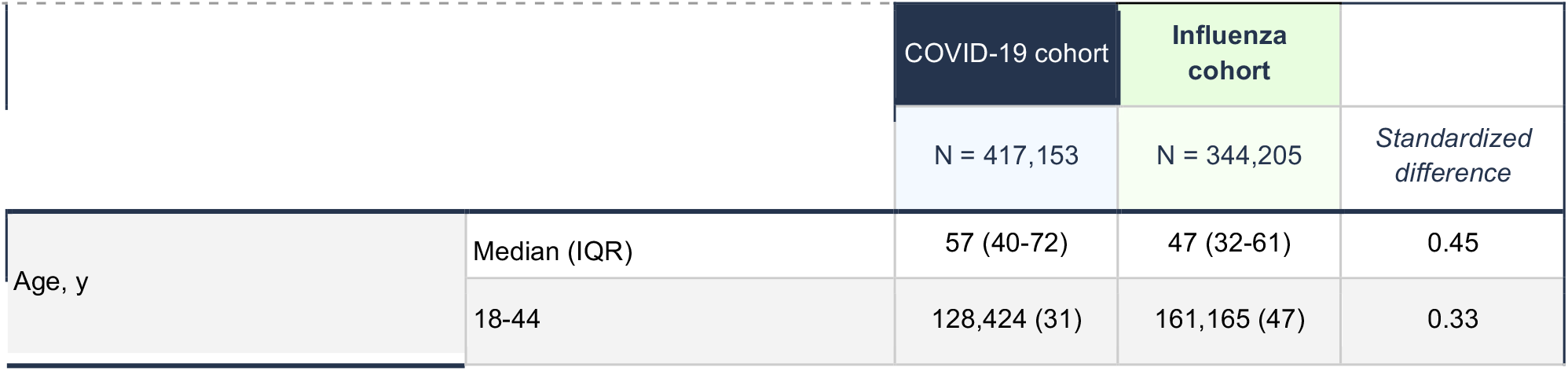
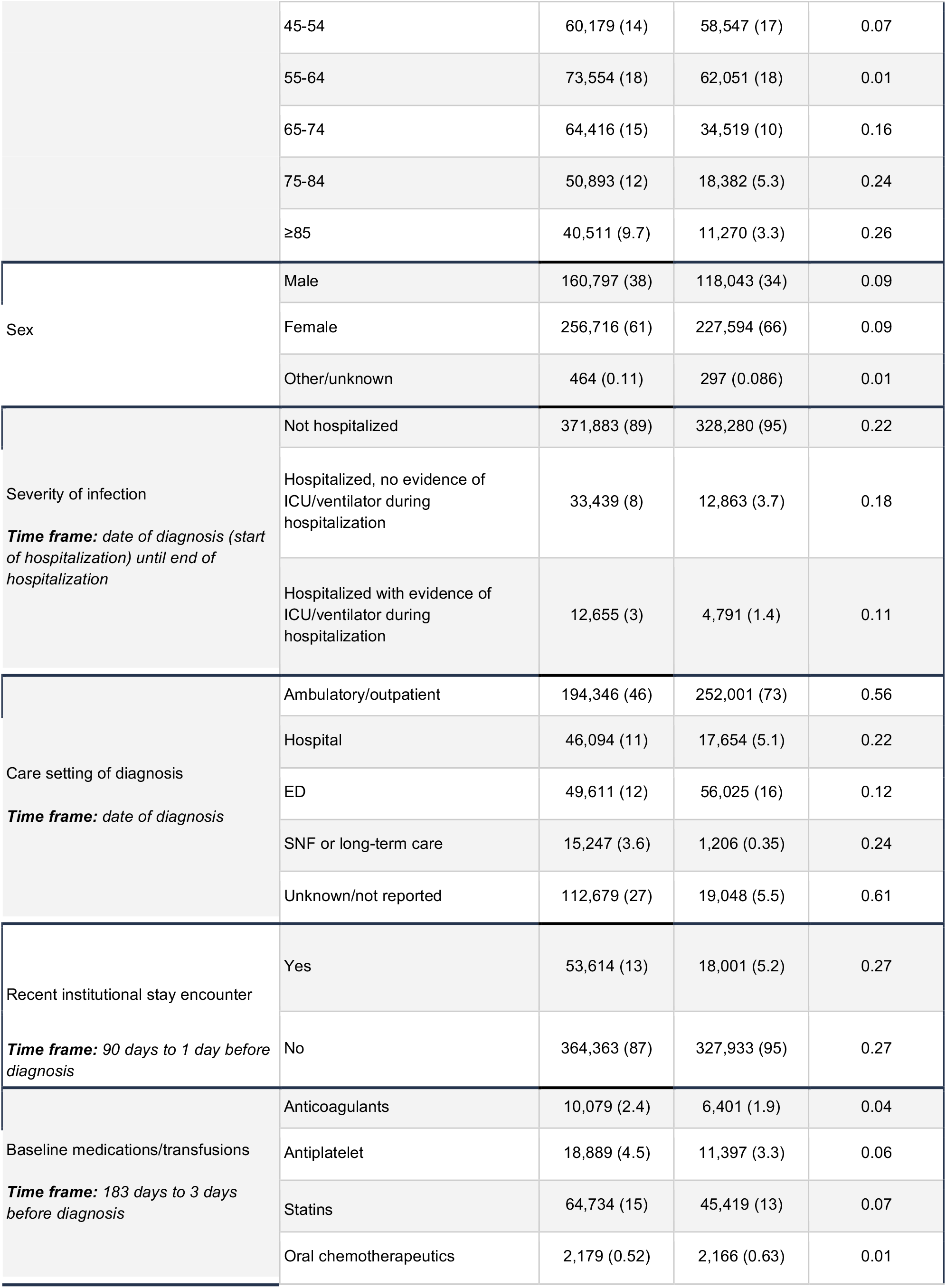
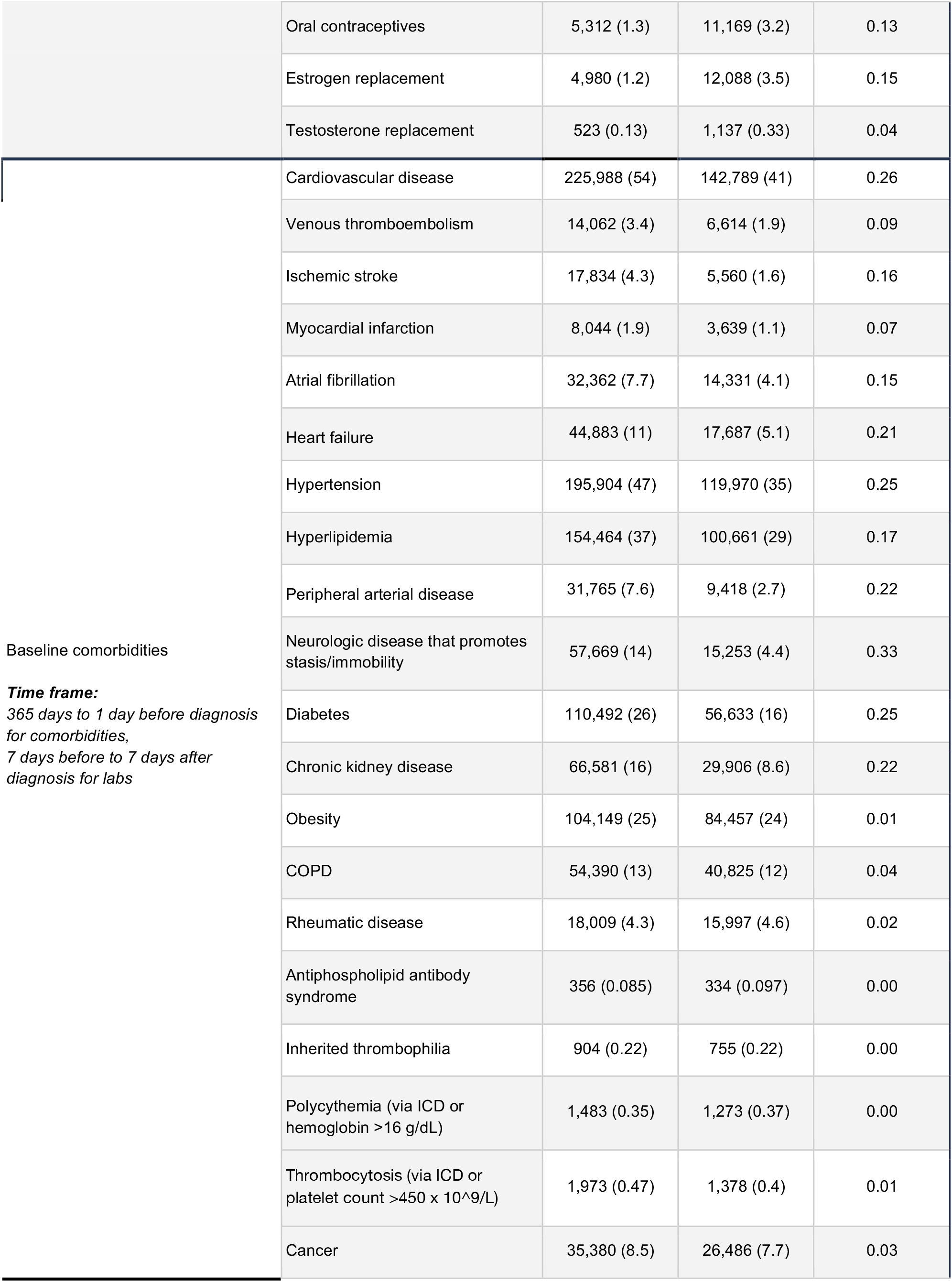
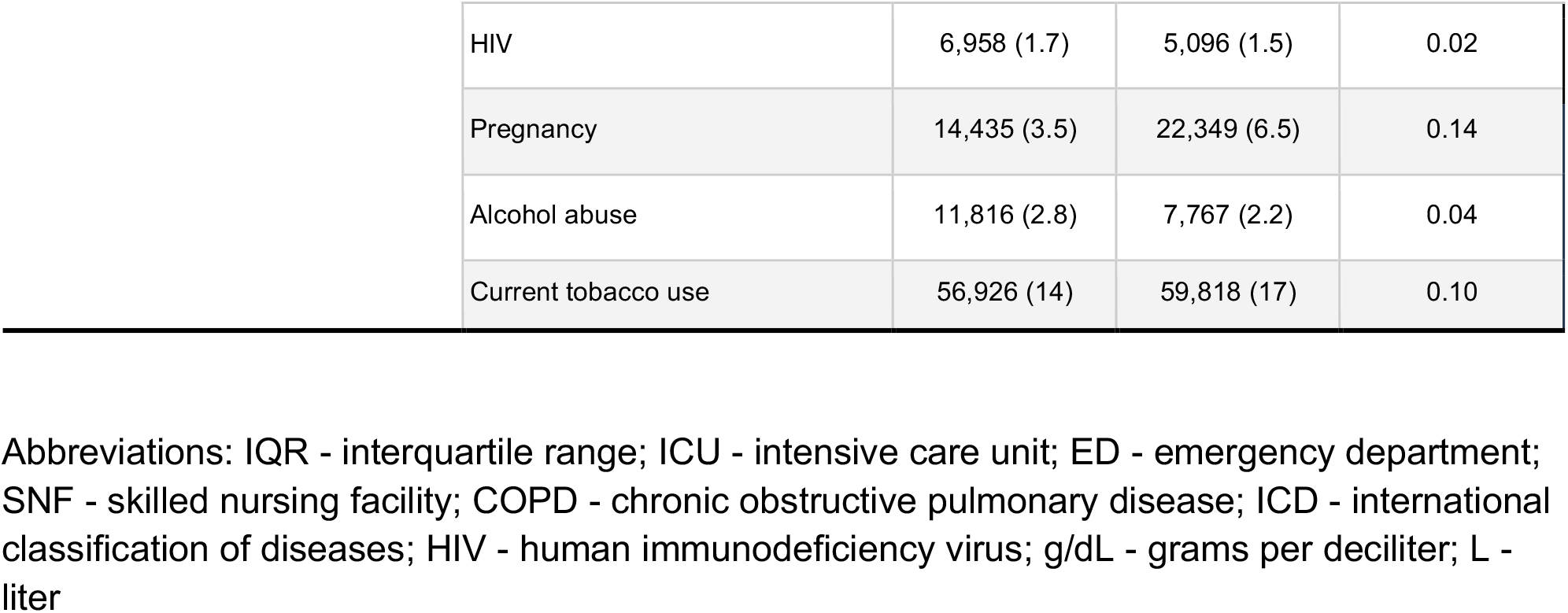
Patient characteristics. Unless otherwise specified, all values are presented as “N (%)”.

Overall, compared to the influenza cohort, the COVID-19 cohort was older, more likely to be male, more likely to have been hospitalized before, at the time of, and after diagnosis, and more likely to have chronic comorbidities. After cohort balancing using the propensity score, all standardized differences between the COVID-19 and influenza cohorts were <0.02 (Supplementary Table 3).

### Primary outcomes

Compared with the influenza cohort, the unadjusted incidence of the primary composite arterial endpoint was more than two-fold higher in the COVID-19 cohort (0.04 versus 0.11 per person-year, respectively, Table 2). However, after adjusting for cohort differences via the propensity score, there was no evidence of increased risk for the primary composite arterial endpoint in COVID-19 patients compared with influenza patients (HR 1.02, 95% CI 0.95 – 1.10, Figure 2).

**Table 2:** Absolute risk and unadjusted incidence rates for primary and secondary outcomes for both cohorts.

|  | COVID-19 cohort |  | Influenza cohort |  |
| --- | --- | --- | --- | --- |
|  | Absolute risk, N (%) | Incidence rates (per person-year) | Absolute risk, N (%) | Incidence rates (per person-year) |
| **Primary Endpoints (Hospital Discharge ICD-10-CM Diagnosis)** |  |  |  |  |
| Arterial thrombosis (combined) | 7,121 (1.7) | 0.11 | 3,035 (0.88) | 0.04 |
| Venous thromboembolism (combined) | 4,278 (1) | 0.06 | 1,524 (0.44) | 0.02 |
| **Secondary Endpoints (ED or hospital discharge ICD-10-CM diagnosis)** |  |  |  |  |
| Acute MI | 4,094 (0.98) | 0.06 | 2,008 (0.58) | 0.03 |
| Acute ischemic or embolic stroke | 3,486 (0.83) | 0.05 | 1,217 (0.35) | 0.02 |
| Acute upper/lower deep venous thrombosis | 2,455 (0.59) | 0.04 | 959 (0.28) | 0.01 |
| Acute pulmonary embolism | 2,315 (0.55) | 0.03 | 772 (0.22) | 0.01 |
| Expanded arterial thrombosis (combined) | 14,462 (3.5) | 0.22 | 7,609 (2.2) | 0.10 |
| Expanded venous thromboembolism (combined) | 4,519 (1.1) | 0.07 | 1,640 (0.47) | 0.02 |

Abbreviations: ICD-10-CM - international classification of diseases, 10th revision; ED - emergency department; MI - myocardial infarction

**Figure 2:**
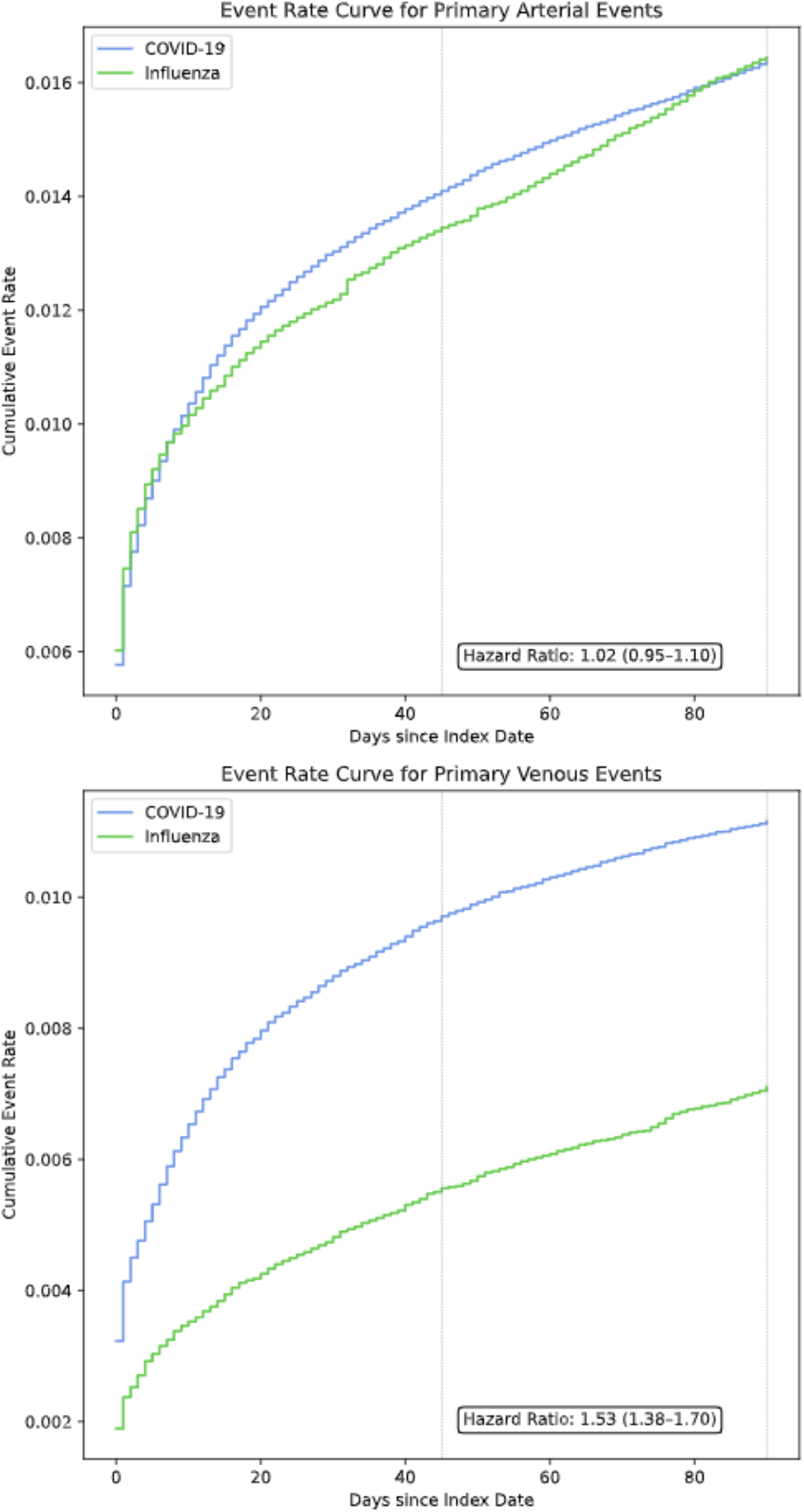
Cumulative event rate curves for primary outcomes. Stratified propensity weighted-cumulative event rate curves in the COVID-19 and influenza populations. After weighting, cohorts were balanced across 49 covariates including demographics, medication use, and clinical comorbidities associated with arterial and venous thromboembolism.

Compared with the influenza cohort, the unadjusted incidence of the primary combined venous endpoint was approximately three-fold higher in the COVID-19 cohort (0.02 versus 0.06 per person-year, respectively, Table 2). After adjusting for cohort differences via the propensity score, there remained a significantly increased risk of the primary venous endpoint in COVID-19 patients compared with influenza patients (HR 1.53, 95% CI 1.38 – 1.70, Figure 2).

### Secondary Outcomes

Stroke and MI demonstrated higher unadjusted incidence rates among COVID-19 patients compared with influenza patients (Table 2). However, upon adjustment for cohort differences via the propensity score, there was no evidence of a difference in risk of stroke (HR 1.11, 95% CI 0.98 – 1.25) or MI (HR 0.93, 95% CI 0.85 – 1.03) between the two cohorts (Figure 3).

**Figure 3:**
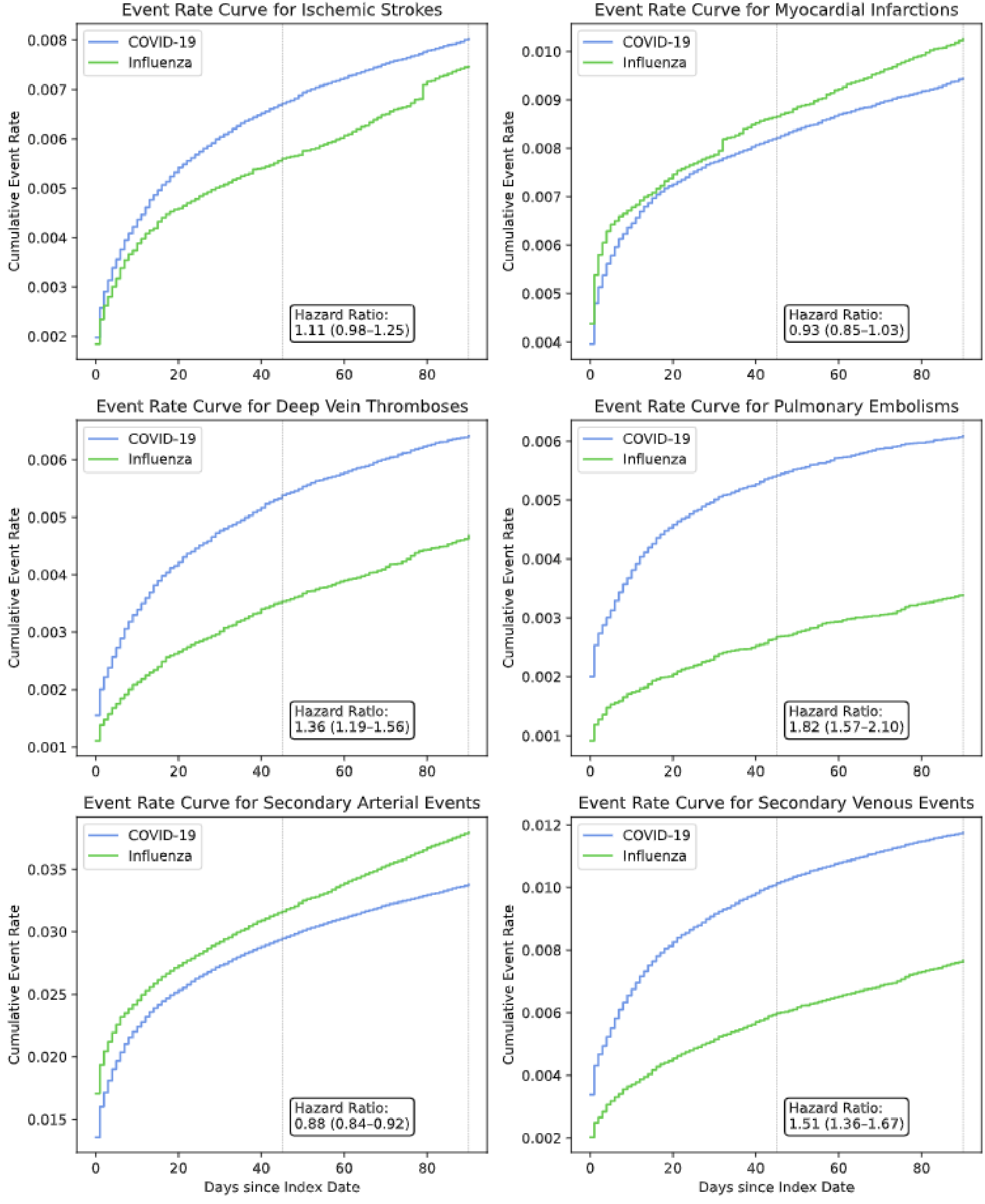
Propensity-weighted cumulative event rate curves for secondary outcomes.

COVID-19 patients demonstrated higher unadjusted incidence rates for DVT and PE compared with influenza patients (Table 2). Upon adjustment for cohort differences via the propensity score, there remained a significantly increased risk for the individual venous outcomes in COVID-19 patients compared with influenza patients (DVT, HR 1.36, 95% CI 1.19 – 1.56; PE, HR 1.82, 95% CI 1.57 – 2.10, Figure 3).

### Sensitivity Analyses

Among patients without prior cardiovascular disease, patients with COVID-19 had a higher risk of the composite arterial endpoint compared with influenza patients (HR 1.46, 95% CI 1.25 – 1.71), while no evidence of a significant difference was seen in patients with prior cardiovascular disease (Table 3).

**Table 3:** COVID-19 vs. influenza hazard ratios for sensitivity cohorts. Hazard ratios (with 95% confidence intervals) for sensitivity cohorts, representing the ratio of outcomes in the COVID-19 cohort as compared to the influenza cohort. Each cohort was separately balanced via propensity score stratification, and the stratified weight was adjusted for in the proportional hazards analysis.

|  | Patients with prior CVD | Patients without prior CVD | Patients with prior VTE | Patients without prior VTE | Inpatients | Outpatients | All patients - unweighted |
| --- | --- | --- | --- | --- | --- | --- | --- |
| Primary arterial endpoint | 0.95 (0.90–1.01) | 1.46 (1.25–1.71) | NA | NA | 0.95 (0.90–1.01) | 1.16 (0.99–1.37) | 2.25 (2.10–2.41) |
| Primary venous endpoint | NA | NA | 1.00 (0.83–1.20) | 1.63 (1.46–1.82) | 1.36 (1.25–1.47) | 1.69 (1.40–2.04) | 2.71 (2.47–2.97) |
| Ischemic stroke | 1.06 (0.97–1.16) | 1.35 (1.07–1.71) | NA | NA | 1.14 (1.04–1.24) | 1.07 (0.83–1.37) | 2.73 (2.46–3.02) |
| Myocardial infarction | 0.87 (0.81–0.93) | 1.50 (1.22–1.84) | NA | NA | 0.85 (0.80–0.91) | 1.23 (1.00–1.52) | 1.95 (1.79–2.12) |
| Deep vein thrombosis | NA | NA | 0.96 (0.79–1.18) | 1.47 (1.27–1.71) | 1.22 (1.09–1.35) | 1.48 (1.18–1.88) | 2.57 (2.29–2.89) |
| Pulmonary embolism | NA | NA | 1.07 (0.78–1.46) | 1.87 (1.61–2.18) | 1.47 (1.31–1.64) | 2.01 (1.54–2.64) | 2.90 (2.56–3.30) |
| Secondary arterial endpoint | 0.84 (0.81–0.87) | 1.42 (1.28–1.59) | NA | NA | 0.84 (0.80–0.87) | 1.01 (0.91–1.11) | 1.76 (1.69–1.84) |
| Secondary venous endpoint | NA | NA | 0.97 (0.81–1.16) | 1.61 (1.45–1.79) | 1.32 (1.22–1.43) | 1.70 (1.42–2.05) | 2.67 (2.45–2.91) |

Abbreviations: CVD - cardiovascular disease; VTE - venous thromboembolism; NA - not assessed.

Among individuals without prior VTE, those diagnosed with COVID-19 had a higher risk of the composite venous endpoint compared with influenza patients (HR 1.63, 95% CI 1.46 – 1.82), while no evidence of a significant difference was seen in patients with prior VTE.

Stratification by inpatient versus outpatient care setting of diagnosis revealed consistent findings across care settings of a similar risk of the composite arterial endpoint and an increased risk of the composite venous endpoint in COVID-19 patients compared with influenza patients.

## DISCUSSION

In a large US cohort linking EHR and claims data, we found that patients with a diagnosis of COVID-19 were at higher adjusted risk for venous thromboembolic events, but not arterial thromboembolic events, compared with patients with influenza. Venous thromboembolic risk was driven by both acute DVT and acute PE events. Upon stratification by prior CVD, a diagnosis of COVID-19 was associated with higher arterial thromboembolic risk in those without prior CVD, but not those with prior CVD. Upon stratification by prior VTE history, a diagnosis of COVID-19 was associated with higher venous thromboembolic risk in those without prior VTE, but not those with prior VTE. Together, these findings address key knowledge gaps regarding the independent thromboembolic risks of COVID-19 compared with influenza and may guide future work to clarify the role of thromboprophylaxis in COVID-19 management.

During the early period of the pandemic, a self-controlled case-series and matched cohort analysis from Sweden suggested that COVID-19 was a risk factor for acute myocardial infarction and ischemic stroke [8]. A single-arm cohort study from Europe indicated a high prevalence of pulmonary embolism among hospitalized COVID-19 patients [9]. Similarly, a single-arm cohort study of 3334 patients in New York City hospitals suggested a high prevalence of 16% for thromboembolic events in COVID-19 patients [10]. These analyses were limited by largely case series-based or single-arm designs. A retrospective cohort study of New York City patients in the emergency department or hospitalized found higher rates of ischemic stroke in patients with COVID-19 compared with those with influenza [11]. However, this study included only selected baseline demographics in the model and did not adjust the results broadly for baseline cohort characteristics that may serve as confounders due to inherent differences between COVID-19 and influenza cohorts. For instance, in our study, COVID-19 patients were more likely to have potentially relevant comorbidities including prior neurologic diseases, CVD, chronic kidney disease, hypertension, diabetes, and heart failure. Additionally, early studies focused on inpatient populations with COVID-19. To our knowledge, ours is the first and largest cohort study to quantify the independent thromboembolic risk of COVID-19 compared with influenza after adjusting broadly for baseline cohort characteristics in a largely outpatient cohort.

In contrast to prior data, we did not find elevated arterial thromboembolic risk among patients with a diagnosis of COVID-19 compared to those with influenza after accounting for differences in baseline clinical status. These findings may have important implications for the natural history and prognosis of COVID-19. Arterial thromboembolic events such as MI and ischemic stroke are associated with poor outcomes and contribute substantially to worldwide morbidity and mortality [23]; given their clinical risks, early reports of elevated arterial event rates served as a key impetus to explore the role of antithrombotic therapy in COVID-19 management. Our work now indicates an urgent need for robust prospective data to validate the present findings and thus, refine our understanding of the need for aggressive antithrombotic therapy. Indeed, our findings of no elevated risk of arterial thrombosis in COVID-19 are consistent with the results of a recent randomized clinical trial that found no evidence of thrombosis reduction benefit with intermediate dose prophylactic anticoagulation compared with standard dose prophylactic anticoagulation in patients with COVID-19 admitted to the intensive care unit [4]. Similarly, a randomized trial of critically ill patients with COVID-19 who were assigned to initial therapeutic dose anticoagulation with heparin versus usual pharmacologic thromboprophylaxis was stopped after meeting the trial’s pre-specified criterion for futility [7]. In addition, an exploratory analysis in our study by prior CVD history suggested that patients without prior CVD experienced higher arterial risk with a diagnosis of COVID-19 compared with influenza. Together, these data lay the foundation to clarify the precise nature of the arterial thromboembolic risk of COVID-19. Especially given a contemporary rise in COVID-19 cases in 2021, addressing this knowledge gap is necessary to develop appropriate thromboprophylaxis strategies for management, and thus, it was a focus for the COVID-19 Evidence Accelerator convened by the Reagan-Udall Foundation for the US FDA.

Our findings validate prior literature supporting elevated venous thromboembolic risk in patients with COVID-19 compared with influenza. These findings support close monitoring for venous events after a diagnosis of COVID-19, as well as the use of strategies to mitigate venous risk including nonpharmacological and pharmacologic prophylaxis [24]. Prior history of VTE may affect this risk, as an exploratory analysis suggested elevated risk in those without prior VTE, but not those with prior VTE. Prospective validation of these findings is warranted.

Our study has additional strengths. We employed a large cohort and linked EHR and claims data to comprehensively capture patient variables and outcomes. To eliminate the possibility of concomitant COVID-19 infection, we selected the comparison cohort of influenza patients from the 2018 influenza season. We adjusted broadly for baseline characteristics with propensity scores to account for cohort differences. We assessed a range of secondary outcomes including individual arterial and venous events as well as composite outcomes. For DVTs, which may be treated in the emergency department setting without hospitalization, we included DVT diagnoses from the ED setting in our outcomes. We performed sensitivity analyses including across care settings (inpatient versus outpatient care), which demonstrated consistent findings.

Our study should be interpreted in the context of certain limitations. Participant-level race/ethnicity data were unavailable, preventing assessment of diverse patient representation and generalizability. Our cohort, although large, may not reflect populations across the United Status. As is inherent to observational data, unmeasured confounding may be present despite efforts to account for baseline characteristics. However, we aimed to characterize the potential impact of unmeasured confounding in an E-value analysis as shown in the Supplement. Death information was unavailable, so we were unable to directly treat death as a censoring event.

However, we included the end of an individual’s claims record (i.e., the last day of a recorded claim or the last day of insurance enrollment) as a censoring event for follow-up - which should typically precede a death event - to minimize the possibility of missing a death event in an included patient.

In conclusion, in a large retrospective cohort linking EHR and claims data of over 750,000 patients, we found that COVID-19 was independently associated with higher 90-day risk for venous thrombosis, but not arterial thrombosis, compared with influenza. Our work warrants expedited prospective validation to help clarify the precise nature of arterial and venous thromboembolic risks in COVID-19 and thus, the role of thromboprophylaxis in COVID-19 management.

## Data Availability

Data used contains de-identified patient-level data and cannot be shared publicly.

## ACKNOWLEDGEMENTS

We would like to acknowledge the Sentinel Initiative, whose COVID-19 Coagulopathy Study Synopsis formed the basis for this study’s methodology. We would also like to acknowledge the Reagan-Udall Foundation for the FDA and the Friends of Cancer Research, who convened the COVID-19 Evidence Accelerator and allowed us to collaborate and refine ideas with other study teams working toward the same research objectives. Further, we would like to thank all of the members of the COVID-19 Natural History of Coagulopathy Parallel Analysis group within the evidence accelerator, including individuals and teams from the Sentinel Initiative, Datavant, Syapse, PCORNet, and the FDA for helpful conversations and valuable feedback.

## Author Contributions

AW contributed to study conceptualization, data curation, formal analysis, investigation, methodology, project administration, software, visualization, manuscript draft preparation and review/editing;

AS contributed to study methodology, manuscript draft preparation and review/editing;

DL contributed to data curation, investigation, software, and manuscript review/editing;

KB contributed to study conceptualization, data curation, investigation, methodology, and manuscript review/editing;

SG contributed to data curation, project administration, and manuscript review/editing;

RB contributed to data curation, investigation, software, project administration and manuscript review/editing;

SC contributed to data curation, investigation, and manuscript review/editing;

MB contributed to data curation, investigation, and manuscript review/editing;

FR contributed to investigation, supervision, and manuscript review/editing;

RD contributed to investigation, methodology, supervision, and manuscript review/editing.

## References

1. Piroth L, Cottenet J, Mariet A-S, Bonniaud P, Blot M, Tubert-Bitter P, et al. Comparison of the characteristics, morbidity, and mortality of COVID-19 and seasonal influenza: a nationwide, population-based retrospective cohort study. Lancet Respir Med. 2021;9: 251– 259.

2. Petersen E, Koopmans M, Go U, Hamer DH, Petrosillo N, Castelli F, et al. Comparing SARS-CoV-2 with SARS-CoV and influenza pandemics. Lancet Infect Dis. 2020;20: e238– e244.

3. Cheruiyot I, Kipkorir V, Ngure B, Misiani M, Munguti J, Ogeng’o J. Arterial Thrombosis in Coronavirus Disease 2019 Patients: A Rapid Systematic Review. Ann Vasc Surg. 2021;70: 273–281.

4. INSPIRATION Investigators, Sadeghipour P, Talasaz AH, Rashidi F, Sharif-Kashani B, Beigmohammadi MT, et al. Effect of Intermediate-Dose vs Standard-Dose Prophylactic Anticoagulation on Thrombotic Events, Extracorporeal Membrane Oxygenation Treatment, or Mortality Among Patients With COVID-19 Admitted to the Intensive Care Unit: The INSPIRATION Randomized Clinical Trial. JAMA. 2021;325: 1620–1630.

5. Al-Samkari H. Finding the Optimal Thromboprophylaxis Dose in Patients With COVID-19. JAMA: the journal of the American Medical Association. 2021. pp. 1613–1615.

6. Paranjpe I, Fuster V, Lala A, Russak AJ, Glicksberg BS, Levin MA, et al. Association of Treatment Dose Anticoagulation With In-Hospital Survival Among Hospitalized Patients With COVID-19. J Am Coll Cardiol. 2020;76: 122–124.

7. REMAP-CAP Investigators, ACTIV-4a Investigators, ATTACC Investigators, Goligher EC, Bradbury CA, McVerry BJ, et al. Therapeutic Anticoagulation with Heparin in Critically Ill Patients with Covid-19. N Engl J Med. 2021;385: 777–789.

8. Katsoularis I, Fonseca-Rodríguez O, Farrington P, Lindmark K, Fors Connolly A-M. Risk of acute myocardial infarction and ischaemic stroke following COVID-19 in Sweden: a self-controlled case series and matched cohort study. Lancet. 2021;398: 599–607.

9. Jevnikar M, Sanchez O, Chocron R, Andronikof M, Raphael M, Meyrignac O, et al. Prevalence of pulmonary embolism in patients with COVID-19 at the time of hospital admission. Eur Respir J. 2021;58. doi:10.1183/13993003.00116-2021

10. Bilaloglu S, Aphinyanaphongs Y, Jones S, Iturrate E, Hochman J, Berger JS. Thrombosis in Hospitalized Patients With COVID-19 in a New York City Health System. JAMA. 2020;324: 799–801.

11. Merkler AE, Parikh NS, Mir S, Gupta A, Kamel H, Lin E, et al. Risk of Ischemic Stroke in Patients With Coronavirus Disease 2019 (COVID-19) vs Patients With Influenza. JAMA Neurol. 2020. doi:10.1001/jamaneurol.2020.2730

12. Lo Re V III, Dutcher SK, Perez-Vilar S, Carbonari DM, Hennessy S, Kempner ME, et al. Study Synopsis: Natural History of Coagulopathy in COVID-19. Sentinel Initiative; 2021 Jun. Report No.: 2. Available: https://www.sentinelinitiative.org/methods-data-tools/methods/assessment-natural-history-coagulopathy-covid-19

13. Lo Re V III, Dutcher SK, Perez-Vilar S, Kit B, Connolly J, Cocoros N. Assessment of the Natural History of Coagulopathy in COVID-19: Statistical Analysis Plan. Sentinel Initiative; 2021 Jul. Report No.: 2. Available: https://www.sentinelinitiative.org/methods-data-tools/methods/assessment-natural-history-coagulopathy-covid-19

14. Stewart M, Rodriguez-Watson C, Albayrak A, Asubonteng J, Belli A, Brown T, et al. COVID-19 Evidence Accelerator: A parallel analysis to describe the use of Hydroxychloroquine with or without Azithromycin among hospitalized COVID-19 patients. PLoS One. 2021;16: e0248128.

15. Katherine Yih W, Hua W, Draper C, Dutcher S, Fuller C, Kempner M, et al. Master protocol development: COVID-19 natural history. 2020 Oct. Available: https://www.sentinelinitiative.org/methods-data-tools/methods/master-protocol-development-covid-19-natural-history

16. Ammann EM, Cuker A, Carnahan RM, Perepu US, Winiecki SK, Schweizer ML, et al. Chart validation of inpatient International Classification of Diseases, Ninth Revision, Clinical Modification (ICD-9-CM) administrative diagnosis codes for venous thromboembolism (VTE) among intravenous immune globulin (IGIV) users in the Sentinel Distributed Database. Medicine. 2018;97: e9960.

17. Ammann EM, Marin L. Schweizer, Robinson JG, Eschol JO, Kafa R, Girotra S, et al. Chart validation of inpatient ICD-9-CM administrative diagnosis codes for acute myocardial infarction (AMI) among intravenous immune globulin (IGIV) users in the Sentinel Distributed Database. Pharmacoepidemiology and Drug Safety. 2018. pp. 398–404. doi:10.1002/pds.4398

18. Cutrona SL, Toh S, Iyer A, Foy S, Daniel GW, Nair VP, et al. Validation of acute myocardial infarction in the Food and Drug Administration’s Mini-Sentinel program. Pharmacoepidemiology and Drug Safety. 2013. pp. 40–54. doi:10.1002/pds.3310

19. Roumie CL, Mitchel E, Gideon PS, Varas-Lorenzo C, Castellsague J, Griffin MR. Validation of ICD-9 codes with a high positive predictive value for incident strokes resulting in hospitalization using Medicaid health data. Pharmacoepidemiology and Drug Safety. 2008. pp. 20–26. doi:10.1002/pds.1518

20. Wahl PM, Rodgers K, Schneeweiss S, Gage BF, Butler J, Wilmer C, et al. Validation of claims-based diagnostic and procedure codes for cardiovascular and gastrointestinal serious adverse events in a commercially-insured population. Pharmacoepidemiol Drug Saf. 2010;19: 596–603.

21. Austin PC. Balance diagnostics for comparing the distribution of baseline covariates between treatment groups in propensity-score matched samples. Stat Med. 2009;28: 3083– 3107.

22. Franklin JM, Eddings W, Austin PC, Stuart EA, Schneeweiss S. Comparing the performance of propensity score methods in healthcare database studies with rare outcomes. Stat Med. 2017;36: 1946–1963.

23. Lozano R, Naghavi M, Foreman K, Lim S, Shibuya K, Aboyans V, et al. Global and regional mortality from 235 causes of death for 20 age groups in 1990 and 2010: a systematic analysis for the Global Burden of Disease Study 2010. Lancet. 2012;380: 2095–2128.

24. Chan NC, Eikelboom JW, Weitz JI. Evolving Treatments for Arterial and Venous Thrombosis: Role of the Direct Oral Anticoagulants. Circ Res. 2016;118: 1409–1424.

