## Supplemental Information for "COVID-19 is associated with higher risk of venous thrombosis, but not arterial thrombosis, compared with influenza: Insights from a large US cohort"

Methods - Additional Details

Since the vast majority of strokes, MIs, and PEs require inpatient treatment, the identification of these endpoints were restricted to inpatient settings to maximize the accuracy of data capture. Inpatient diagnoses were identified as those which were attached to a claim which originated from an inpatient hospital (CMS place of service 21)[1]. However, because a significant percentage of acute DVTs do not require inpatient hospitalization [2], the identification of acute DVT was expanded to the emergency department setting (CMS place of service 23) as well as the inpatient setting. The identification of secondary endpoints (angina, transient ischemic attack, peripheral arterial disease, amputation, coronary angioplasty, coronary artery bypass grafting, venous thrombosis of devices, implants, or grafts) were similarly restricted to ICD codes attached to claims which originated from either an inpatient hospital or emergency room (CMS place of service 21 or 23) [1].

COVID-19 ICD diagnosis codes and LOINC codes for SARS-CoV-2 molecular tests were sourced from the Sentinel Initiative. SARS-CoV-2 antigen tests have been shown to have lower sensitivities and specificities than molecular tests, and were not considered. April 1, 2020 served as the study start date, as that is the date when the ICD code for COVID-19 (U07.1) was released by the CDC; before this, testing was also not widespread.

The baseline period was defined as the 365 days prior to the index date; individuals were only excluded if they have fewer than two encounters in EHR data AND fewer than two claims in insurance claims data within the baseline period (inclusive of the index date). Pediatric patients (under 18 years of age at their index date) were excluded, as the effects of COVID-19 are less pronounced in this population. Patients with no claims information after the index date were excluded to avoid mistakenly presuming patients were healthy after the index date because they have no claims data, when in reality they had experienced outcomes that were not recorded in the dataset (a form of misclassification bias).

Evidence of ICU and/or mechanical ventilation were identified via CPT/HCPCS codes as specified by the Sentinel Initiative.

Propensity score stratification was chosen as the method to balance cohorts in order to efficiently retain individuals in the analysis when handling imbalanced cohort sizes.

Several other propensity score balancing methods were considered, including 1:1 patient matching, inverse probability of treatment weighting (IPTW) with weights stabilized based on cohort size, standardized mortality ratio weighting (SMRW) with weights truncated at 8, and propensity score stratification using 500 bins.

For the individuals with COVID-19, stratified propensity weights ($w_i$) were calculated using the following formula:

$$w_i = \frac{n_b n_{COVID-19}}{n_{b,\quad COVID-19} N}$$

where $n_b$ is the number of patients in bin *b*, $n_{COVID-19}$ is the number of patients with COVID-19 in bin *b*, $n_{b,\quad COVID-19}$ is the number of patients in bin *b* with COVID-19, and *N* is the total number of patients.

For individuals with influenza, stratified propensity weights ($w_i$) were calculated using the following formula:

$$w_i = \frac{n_b n_{influenza}}{n_{b,\quad influenza} N}$$

where $n_{influenza}$ is the number of patients with influenza in bin *b*, and $n_{b,\quad influenza}$ is the number of patients in bin *b* with influenza.

For each sensitivity analysis, propensity scores were recalculated (i.e. a new regression classifier was trained on the sub-cohort included in the sensitivity analysis), and new stratified weights were computed.

To assess the impact of unmeasured confounders, an E-value was calculated for the 8 primary and secondary endpoints. This E-value determines the minimum strength an unmeasured variable would have to be associated with both the exposure (influenza vs. COVID-19) and the outcome in order for there to be no difference in the two cohorts (or, for the value of 1.0 to fall within the 95% confidence interval of the calculated hazard ratio). These results are presented in Supplemental Table 4. Analysis was performed in R version 4.1.1, using the "EValue" package [3].

Supplemental Table 1: ICD codes to define COVID-19 and influenza

| **Diagnosis** | **ICD-10-CM** | **Description** |
|---|---|---|
| COVID-19 | B9729 | Other coronavirus as the cause of diseases classified elsewhere |
| COVID-19 | U071 | COVID-19, virus identified [code effective April 1, 2020] |
| COVID-19 | B342 | Coronavirus infection, unspecified site |

| | | |
|---|---|---|
| COVID-19 | B9721 | SARS-associated coronavirus as the cause of diseases classified elsewhere |
| COVID-19 | J1281 | Pneumonia due to SARS-associated coronavirus |
| Influenza | J09 | Influenza due to certain identified influenza viruses |
| Influenza | J09X | Influenza due to identified novel influenza A virus |
| Influenza | J09X1 | Influenza due to identified novel influenza A virus with pneumonia |
| Influenza | J09X2 | Influenza due to identified novel influenza A virus with other respiratory manifestations |
| Influenza | J09X3 | Influenza due to identified novel influenza A virus with gastrointestinal manifestations |
| Influenza | J09X9 | Influenza due to identified novel influenza A virus with other manifestations |
| Influenza | J10 | Influenza due to other identified influenza virus |
| Influenza | J100 | Influenza due to other identified influenza virus with pneumonia |
| Influenza | J1000 | Influenza due to other identified influenza virus with unspecified type of pneumonia |
| Influenza | J1001 | Influenza due to other identified influenza virus with the same other identified influenza virus pneumonia |
| Influenza | J1008 | Influenza due to other identified influenza virus with other specified pneumonia |
| Influenza | J101 | Influenza due to other identified influenza virus with other respiratory manifestations |
| Influenza | J102 | Influenza due to other identified influenza virus with gastrointestinal manifestations |
| Influenza | J108 | Influenza due to other identified influenza virus with other manifestations |
| Influenza | J1081 | Influenza due to other identified influenza virus with encephalopathy |
| Influenza | J1082 | Influenza due to other identified influenza virus with myocarditis |
| Influenza | J1083 | Influenza due to other identified influenza virus with otitis media |
| Influenza | J1089 | Influenza due to other identified influenza virus with other manifestations |
| Influenza | J11 | Influenza due to unidentified influenza virus |
| Influenza | J110 | Influenza due to unidentified influenza virus with pneumonia |
| Influenza | J1100 | Influenza due to unidentified influenza virus with unspecified type of pneumonia |

| | | |
|---|---|---|
| Influenza | J1108 | Influenza due to unidentified influenza virus with specified pneumonia |
| Influenza | J111 | Influenza due to unidentified influenza virus with other respiratory manifestations |
| Influenza | J112 | Influenza due to unidentified influenza virus with gastrointestinal manifestations |
| Influenza | J118 | Influenza due to unidentified influenza virus with other manifestations |
| Influenza | J1181 | Influenza due to unidentified influenza virus with encephalopathy |
| Influenza | J1182 | Influenza due to unidentified influenza virus with myocarditis |
| Influenza | J1183 | Influenza due to unidentified influenza virus with otitis media |
| Influenza | J1189 | Influenza due to unidentified influenza virus with other manifestations |

Supplemental Table 2: LOINC codes used to identify SARS-COV-2 and influenza molecular tests

| **Diagnosis** | **LOINC** | **Description** |
|---|---|---|
| COVID-19 | 94306-8 | SARS coronavirus 2 RNA panel - Unspecified specimen by NAA with probe detection |
| COVID-19 | 94307-6 | SARS coronavirus 2 N gene [Presence] in Unspecified specimen by Nucleic acid amplification using primer-probe set N1 |
| COVID-19 | 94308-4 | SARS coronavirus 2 N gene [Presence] in Unspecified specimen by Nucleic acid amplification using primer-probe set N2 |
| COVID-19 | 94309-2 | SARS coronavirus 2 RNA [Presence] in Unspecified specimen by NAA with probe detection |
| COVID-19 | 94310-0 | SARS-like coronavirus N gene [Presence] in Unspecified specimen by NAA with probe detection |
| COVID-19 | 94311-8 | SARS coronavirus 2 N gene [Cycle Threshold #] in Unspecified specimen by Nucleic acid amplification using primer-probe set N1 |
| COVID-19 | 94312-6 | SARS coronavirus 2 N gene [Cycle Threshold #] in Unspecified specimen by Nucleic acid amplification using primer-probe set N2 |
| COVID-19 | 94313-4 | SARS-like coronavirus N gene [Cycle Threshold #] in Unspecified specimen by NAA with probe detection |
| COVID-19 | 94314-2 | SARS coronavirus 2 RdRp gene [Presence] in Unspecified specimen by NAA with probe detection |
| COVID-19 | 94315-9 | SARS coronavirus 2 E gene [Presence] in Unspecified specimen by NAA with probe detection |

| | | |
|---|---|---|
| COVID-19 | 94316-7 | SARS coronavirus 2 N gene [Presence] in Unspecified specimen by NAA with probe detection |
| COVID-19 | 94500-6 | SARS coronavirus 2 RNA [Presence] in Respiratory specimen by NAA with probe detection |
| COVID-19 | 94502-2 | SARS-related coronavirus RNA [Presence] in Respiratory specimen by NAA with probe detection |
| COVID-19 | 94509-7 | SARS coronavirus 2 E gene [Cycle Threshold #] in Unspecified specimen by NAA with probe detection |
| COVID-19 | 94510-5 | SARS coronavirus 2 N gene [Cycle Threshold #] in Unspecified specimen by NAA with probe detection |
| COVID-19 | 94511-3 | SARS coronavirus 2 ORF1ab region [Cycle Threshold #] in Unspecified specimen by NAA with probe detection |
| COVID-19 | 94531-1 | SARS coronavirus 2 RNA panel - Respiratory specimen by NAA with probe detection |
| COVID-19 | 94532-9 | SARS-related coronavirus+MERS coronavirus RNA [Presence] in Respiratory specimen by NAA with probe detection |
| COVID-19 | 94533-7 | SARS coronavirus 2 N gene [Presence] in Respiratory specimen by NAA with probe detection |
| COVID-19 | 94534-5 | SARS coronavirus 2 RdRp gene [Presence] in Respiratory specimen by NAA with probe detection |
| COVID-19 | 94559-2 | SARS coronavirus 2 ORF1ab region [Presence] in Respiratory specimen by NAA with probe detection |
| COVID-19 | 94565-9 | SARS coronavirus 2 RNA [Presence] in Nasopharynx by NAA with non-probe detection |
| COVID-19 | 94639-2 | SARS coronavirus 2 ORF1ab region [Presence] in Unspecified specimen by NAA with probe detection |
| COVID-19 | 94640-0 | SARS coronavirus 2 S gene [Presence] in Respiratory specimen by NAA with probe detection |
| COVID-19 | 94641-8 | SARS coronavirus 2 S gene [Presence] in Unspecified specimen by NAA with probe detection |
| COVID-19 | 94642-6 | SARS coronavirus 2 S gene [Cycle Threshold #] in Respiratory specimen by NAA with probe detection |
| COVID-19 | 94643-4 | SARS coronavirus 2 S gene [Cycle Threshold #] in Unspecified specimen by NAA with probe detection |
| COVID-19 | 94644-2 | SARS coronavirus 2 ORF1ab region [Cycle Threshold #] in Respiratory specimen by NAA with probe detection |
| COVID-19 | 94645-9 | SARS coronavirus 2 RdRp gene [Cycle Threshold #] in Unspecified specimen by NAA with probe detection |

| | | |
|---|---|---|
| COVID-19 | 94646-7 | SARS coronavirus 2 RdRp gene [Cycle Threshold #] in Respiratory specimen by NAA with probe detection |
| COVID-19 | 94647-5 | SARS-related coronavirus RNA [Presence] in Unspecified specimen by NAA with probe detection |
| COVID-19 | 94660-8 | SARS coronavirus 2 RNA [Presence] in Serum or Plasma by NAA with probe detection |
| COVID-19 | 94745-7 | SARS coronavirus 2 RNA |
| COVID-19 | 94746-5 | SARS coronavirus 2 RNA |
| COVID-19 | 94756-4 | SARS coronavirus 2 N gene |
| COVID-19 | 94757-2 | SARS coronavirus 2 N gene |
| COVID-19 | 94758-0 | SARS-related coronavirus E gene |
| COVID-19 | 94759-8 | SARS coronavirus 2 RNA |
| COVID-19 | 94760-6 | SARS coronavirus 2 N gene |
| COVID-19 | 94765-5 | SARS-related coronavirus E gene |
| COVID-19 | 94766-3 | SARS coronavirus 2 N gene |
| COVID-19 | 94767-1 | SARS coronavirus 2 S gene |
| COVID-19 | 94819-0 | SARS coronavirus 2 RNA |
| COVID-19 | 94845-5 | SARS coronavirus 2 RNA |
| COVID-19 | 95380-2 | Influenza virus A + B and SARS-CoV-2 (COVID-19) and SARS-related CoV RNA panel - Respiratory specimen by NAA with probe detection |
| COVID-19 | 95406-5 | SARS coronavirus 2 RNA |
| COVID-19 | 95409-9 | SARS coronavirus 2 N gene |
| COVID-19 | 95422-2 | Influenza virus A + B RNA and SARS-CoV-2 (COVID-19) N gene panel - Respiratory specimen by NAA with probe detection |

| | | |
|---|---|---|
| COVID-19 | 95423-0 | Influenza virus A + B and SARS-CoV-2 (COVID-19) identified in Respiratory specimen by NAA with probe detection |
| COVID-19 | 95425-5 | SARS-CoV-2 (COVID-19) N gene [Presence] in Saliva (oral fluid) by NAA with probe detection |
| COVID-19 | 95521-1 | SARS coronavirus 2 N gene |
| COVID-19 | 95522-9 | SARS coronavirus 2 N gene |
| COVID-19 | 95608-6 | SARS coronavirus 2 RNA |
| Influenza | 344879 | Influenza virus A RNA [Presence] in Unspecified specimen by NAA with probe detection |
| Influenza | 409821 | Influenza virus B RNA [Presence] in Unspecified specimen by NAA with probe detection |
| Influenza | 485094 | Influenza virus A and B RNA [Identifier] in Unspecified specimen by NAA with probe detection |
| Influenza | 502195 | Respiratory pathogens DNA and RNA 12a panel - Unspecified specimen by NAA with probe detection |
| Influenza | 299073 | Haemophilus influenzae B DNA [Presence] in Unspecified specimen by NAA with probe detection |
| Influenza | 299099 | Parainfluenza virus 2 RNA [Presence] in Unspecified specimen by NAA with probe detection |
| Influenza | 299107 | Parainfluenza virus 3 RNA [Presence] in Unspecified specimen by NAA with probe detection |
| Influenza | 299065 | Haemophilus influenzae A DNA [Presence] in Unspecified specimen by NAA with probe detection |
| Influenza | 299081 | Parainfluenza virus 1 RNA [Presence] in Unspecified specimen by NAA with probe detection |
| Influenza | 410100 | Parainfluenza virus 4 RNA [Presence] in Unspecified specimen by NAA with probe detection |
| Influenza | 624627 | Influenza virus A+B RNA [Presence] in Unspecified specimen by NAA with probe detection |
| Influenza | 821710 | Parainfluenza virus 1 RNA [Presence] in Nasopharynx by NAA with non-probe detection |
| Influenza | 821660 | Influenza virus A RNA [Presence] in Nasopharynx by NAA with non-probe detection |
| Influenza | 821728 | Parainfluenza virus 2 RNA [Presence] in Nasopharynx by NAA with non-probe detection |

| | | |
|---|---|---|
| Influenza | 821744 | Parainfluenza virus 4 RNA [Presence] in Nasopharynx by NAA with non-probe detection |
| Influenza | 821702 | Influenza virus B RNA [Presence] in Nasopharynx by NAA with non-probe detection |
| Influenza | 821736 | Parainfluenza virus 3 RNA [Presence] in Nasopharynx by NAA with non-probe detection |
| Influenza | 821678 | Influenza virus A H1 RNA [Presence] in Nasopharynx by NAA with non-probe detection |
| Influenza | 495242 | Influenza virus A H3 RNA [Presence] in Unspecified specimen by NAA with probe detection |
| Influenza | 495218 | Influenza virus A H1 RNA [Presence] in Unspecified specimen by NAA with probe detection |
| Influenza | 821694 | Influenza virus A H3 RNA [Presence] in Nasopharynx by NAA with non-probe detection |
| Influenza | 821686 | Influenza virus A H1 2009 pandemic RNA [Presence] in Nasopharynx by NAA with non-probe detection |
| Influenza | 554659 | Influenza virus A H1 2009 pandemic RNA [Presence] in Unspecified specimen by NAA with probe detection |
| Influenza | 554634 | Influenza virus A swine origin RNA [Identifier] in Unspecified specimen by NAA with probe detection |
| Influenza | 760785 | Influenza virus A RNA [Presence] in Nasopharynx by NAA with probe detection |
| Influenza | 760801 | Influenza virus B RNA [Presence] in Nasopharynx by NAA with probe detection |
| Influenza | 770289 | Influenza virus A H1 2009 pandemic RNA [Presence] in Nasopharynx by NAA with probe detection |
| Influenza | 805903 | Influenza virus A H3 HA gene [Presence] in Nasopharynx by NAA with probe detection |
| Influenza | 770263 | Influenza virus A H1 RNA [Presence] in Nasopharynx by NAA with probe detection |
| Influenza | 770271 | Influenza virus A H3 RNA [Presence] in Nasopharynx by NAA with probe detection |
| Influenza | 495317 | Influenza virus A RNA [Presence] in Isolate by NAA with probe detection |
| Influenza | 613661 | Haemophilus influenzae DNA [Presence] in Unspecified specimen by NAA with probe detection |
| Influenza | 495358 | Influenza virus B RNA [Presence] in Isolate by NAA with probe detection |

Supplemental Table 3: Standardized differences of cohort characteristics before and after weighting, for the primary (full) cohort. Standardized differences for sensitivity cohorts are available on request.

| | | COVID-19 cohort | Influenza cohort | | |
|---|---|---|---|---|---|
| | | N = 417,969 (after trimming the tails) | N = 345,934 (after trimming the tails) | *Standardized difference (before weighting)* | *Standardized difference (after weighting)* |
| Age | 18-44 | 128,424 (31) | 161,165 (47) | 0.33 | 0.011 |
| | 45-54 | 60,179 (14) | 58,547 (17) | 0.07 | 0.006 |
| | 55-64 | 73,553 (18) | 62,051 (18) | 0.01 | 0.010 |
| | 65-74 | 64,416 (15) | 34,519 (10) | 0.16 | 0.010 |
| | 75-84 | 50,892 (12) | 18,382 (5.3) | 0.24 | 0.000 |
| | ≥85 | 40,505 (9.7) | 11,270 (3.3) | 0.26 | 0.015 |
| Sex | Male | 160,795 (38) | 118,043 (34) | 0.09 | 0.007 |
| | Female | 256,710 (61) | 227,594 (66) | 0.09 | 0.007 |
| | Other/unknown | 464 (0.11) | 297 (0.086) | 0.01 | 0.008 |
| Severity of infection<br><br>***Time frame:*** *date of diagnosis (start of hospitalization) until end of hospitalization* | Not hospitalized | 371,878 (89) | 328,280 (95) | 0.22 | 0.079 |
| | Hospitalized, no evidence of ICU/ventilator during hospitalization | 33,437 (8) | 12,863 (3.7) | 0.18 | 0.065 |
| | Hospitalized with evidence of ICU/ventilator during hospitalization | 12,654 (3) | 4,791 (1.4) | 0.11 | 0.041 |
| Care setting of diagnosis | Ambulatory/outpatient | 194,346 (46) | 252,001 (73) | 0.56 | 0.359 |

| | | | | | |
|---|---|---|---|---|---|
| ***Time frame:*** *date of diagnosis* | Hospital | 46,091 (11) | 17,654 (5.1) | 0.22 | 0.079 |
| | ED | 49,611 (12) | 56,025 (16) | 0.12 | 0.103 |
| | SNF or long-term care | 15,244 (3.6) | 1,206 (0.35) | 0.24 | 0.120 |
| | Unknown/not reported | 112,677 (27) | 19,048 (5.5) | 0.61 | 0.483 |
| Recent institutional stay encounter (90 - 1 day before index)<br><br>***Time frame:*** *90 - 1 day before diagnosis* | Yes | 53,607 (13) | 18,001 (5.2) | 0.27 | 0.003 |
| | No | 364,362 (87) | 327,933 (95) | 0.27 | 0.003 |
| Baseline medications/transfusions<br><br>***Time frame:*** *183 - 3 days before diagnosis* | Anticoagulants | 10,079 (2.4) | 6,401 (1.9) | 0.04 | 0.007 |
| | Antiplatelet | 18,887 (4.5) | 11,397 (3.3) | 0.06 | 0.012 |
| | Statins | 64,733 (15) | 45,419 (13) | 0.07 | 0.010 |
| | Oral chemotherapeutics | 5,312 (1.3) | 11,169 (3.2) | 0.13 | 0.007 |
| | Tamoxifen | 2,179 (0.52) | 2,166 (0.63) | 0.01 | 0.002 |
| | Oral contraceptives | 4,979 (1.2) | 12,088 (3.5) | 0.15 | 0.010 |
| | Estrogen replacement | 58 (0.014) | 146 (0.042) | 0.02 | 0.002 |
| | Testosterone replacement | 522 (0.12) | 1,137 (0.33) | 0.04 | 0.003 |
| Baseline comorbidities<br><br>***Time frame:***<br>*365 - 1 day before diagnosis for comorbidities,*<br>*7 days before - 7 days after diagnosis for labs* | Cardiovascular disease | 225,980 (54) | 142,789 (41) | 0.26 | 0.001 |
| | Venous thromboembolism | 14,056 (3.4) | 6,614 (1.9) | 0.09 | 0.008 |
| | Neurologic disease that promotes stasis/immobility | 57,662 (14) | 15,253 (4.4) | 0.33 | 0.001 |
| | Obesity | 104,144 (25) | 84,457 (24) | 0.01 | 0.011 |
| | Alcohol abuse | 11,816 (2.8) | 7,767 (2.2) | 0.04 | 0.000 |

| | | | | |
|---|---|---|---|---|
| Current tobacco use | 56,925 (14) | 59,818 (17) | 0.10 | 0.009 |
| Pregnancy | 14,435 (3.5) | 22,349 (6.5) | 0.14 | 0.008 |
| Chronic kidney disease | 66,574 (16) | 29,906 (8.6) | 0.22 | 0.016 |
| cancer | 35,379 (8.5) | 26,486 (7.7) | 0.03 | 0.005 |
| COPD | 54,389 (13) | 40,825 (12) | 0.04 | 0.007 |
| diabetes | 110,485 (26) | 56,633 (16) | 0.25 | 0.010 |
| hyperlipidemia | 154,459 (37) | 100,661 (29) | 0.17 | 0.009 |
| hypertension | 195,897 (47) | 119,970 (35) | 0.25 | 0.004 |
| rheumatic disease | 18,009 (4.3) | 15,997 (4.6) | 0.02 | 0.006 |
| atrial fibrillation | 32,360 (7.7) | 14,331 (4.1) | 0.15 | 0.010 |
| antiphospholipid antibody syndrome | 356 (0.085) | 334 (0.097) | 0.00 | 0.000 |
| inherited thrombophilia | 904 (0.22) | 755 (0.22) | 0.00 | 0.003 |
| ischemic stroke | 17,827 (4.3) | 5,560 (1.6) | 0.16 | 0.011 |
| myocardial infarction | 8,042 (1.9) | 3,639 (1.1) | 0.07 | 0.009 |
| heart failure | 44,876 (11) | 17,687 (5.1) | 0.21 | 0.021 |
| peripheral arterial disease | 31,758 (7.6) | 9,418 (2.7) | 0.22 | 0.010 |
| Polycythemia (via ICD or hemoglobin >16 g/dL) | 1,483 (0.35) | 1,273 (0.37) | 0.00 | 0.001 |
| Thrombocytosis (via ICD or platelet count >450 x 10^9/L) | 1,973 (0.47) | 1,378 (0.4) | 0.01 | 0.000 |

Supplemental Table 4: Unadjusted event rates for components of secondary composite endpoints

| NOTE 1: this table should be reproduced in the three secondary analysis cohorts (inpatient diagnoses, outpatient diagnoses, NAAT diagnoses) | | COVID-19 cohort | | Influenza cohort | |
|---|---|---|---|---|---|
| | | Absolute risk (N, %) | Incidence rates (per person-year) | Absolute risk (N, %) | Incidence rates (per person-year) |
| | Angina | 1,692 (0.4) | 0.03 | 1,353 (0.39) | 0.02 |
| | Peripheral arterial disease | 2,917 (0.7) | 0.04 | 1,306 (0.38) | 0.02 |
| | Amputation | 1,321 (0.32) | 0.02 | 550 (0.16) | 0.01 |
| | Coronary angioplasty | 2,760 (0.66) | 0.04 | 2,517 (0.73) | 0.03 |
| | Coronary artery bypass grafting | 4,690 (1.1) | 0.07 | 3,428 (0.99) | 0.04 |
| | Venous thrombosis of devices, implants, or grafts | 309 (0.074) | 0.00 | 153 (0.044) | 0.00 |

Supplemental Table 5: E-values (point estimate and 95% confidence intervals) for primary and secondary outcomes. NOTE: for endpoints for which hazard ratios were not statistically significant, no E-values were calculated.

| | E value, point estimate (95% CI) |
|---|---|
| Primary arterial endpoint | NA |
| Primary venous endpoint | 2.43 (2.1) |
| Ischemic stroke | NA |
| Myocardial infarction | NA |
| Deep vein thrombosis | 2.06 (1.67) |
| Pulmonary embolism | 3.04 (2.52) |
| Secondary arterial endpoint | 1.53 (1.39) |

| | |
|---|---|
| Secondary venous endpoint | 2.39 (2.06) |

Supplemental Figure 1: standardized difference, visualized, for different propensity score methods (before and after balancing).
Abbreviations: IPTW - inverse probability of treatment weighting; SMRW - standardized mortality ratio weighting

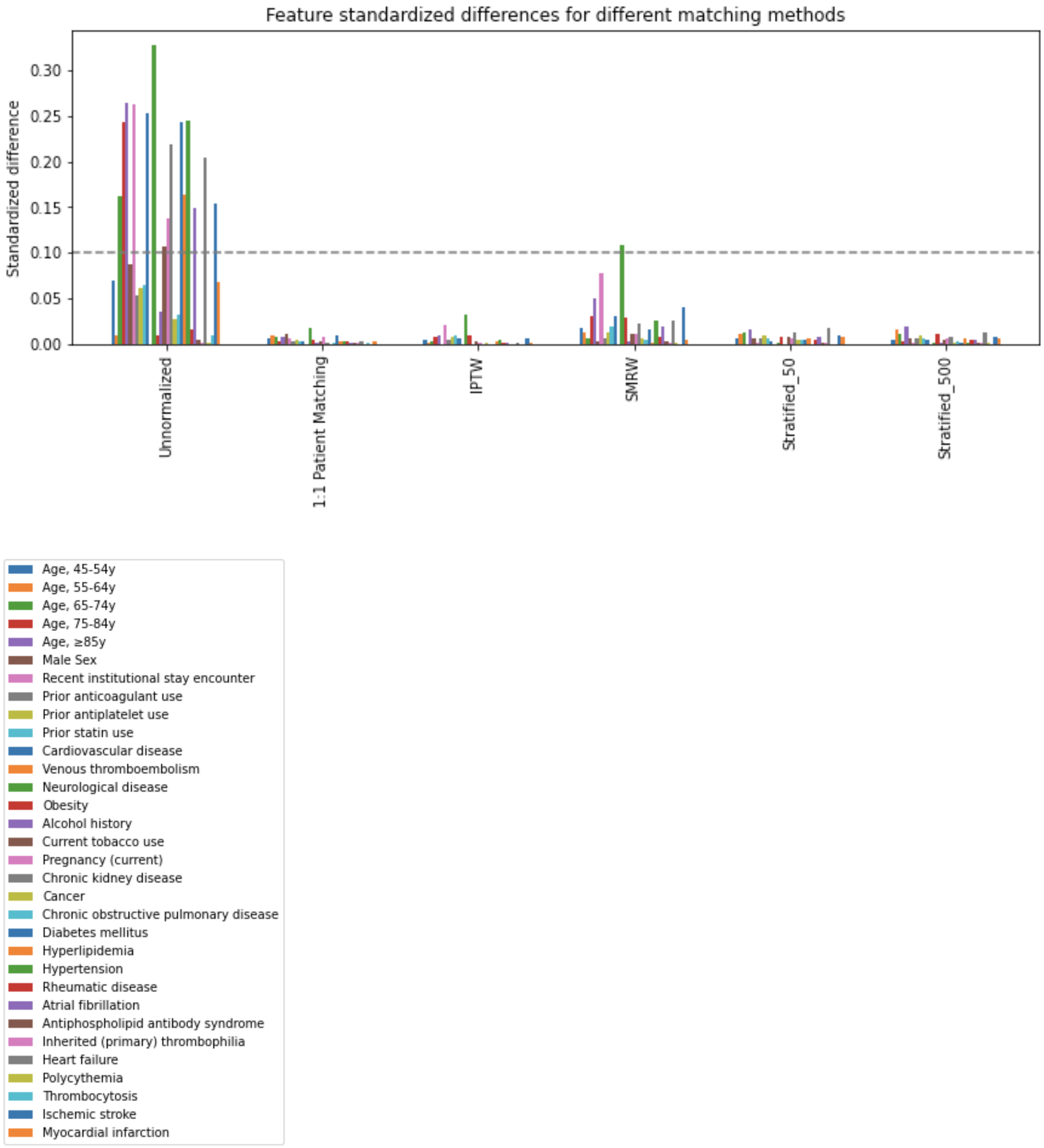


### References


1. Place of Service Code Set. [cited 20 Aug 2021]. Available: https://www.cms.gov/Medicare/Coding/place-of-service-codes/Place_of_Service_Code_Set

2. Dentali F, Di Micco G, Giorgi Pierfranceschi M, Gussoni G, Barillari G, Amitrano M, et al.

Rate and duration of hospitalization for deep vein thrombosis and pulmonary embolism in real-world clinical practice. Ann Med. 2015;47: 546–554.

3. Mathur MB, Ding P, Riddell CA, VanderWeele TJ. Web Site and R Package for Computing E-values. Epidemiology. 2018;29: e45–e47.
